# Functional brain changes after pharmacological interventions in post-traumatic stress disorder: A systematic review of clinical trials

**DOI:** 10.1101/2023.02.28.23286608

**Authors:** Shahab Lotfinia, Amin Afshar, Aram Yaseri, Miranda Olff, Yann Quidé

## Abstract

**Background:** Posttraumatic stress disorder (PTSD) is a complex and heterogeneous mental health condition that can develop after exposure to a traumatic event. Clinical trials have used new pharmacological agents to treat PTSD, but their associated neural correlates remain unclear. The present systematic review aims to summarise the changes in brain function associated with the use these pharmacological agents in PTSD.

**Methods:** Clinical trials using functional magnetic resonance imaging (fMRI), either at rest or during the performance of tasks, were included if they compared the effects of pharmacological agents in patients with PTSD to either trauma-exposed controls or never exposed to trauma healthy controls.

**Results:** Twelve studies were included, of which eight used intranasal oxytocin, two used hydrocortisone, and one used Tolcapone. Oxytocin administration was associated with normalisation of functional connectivity between the ventromedial prefrontal cortex and amygdala, as well as enhanced the function of brain regions specifically involved in emotion processing (e.g., amygdala), working memory (e.g., dorsolateral prefrontal cortex), reward (putamen). Hydrocortisone did not influence brain function at rest or during the performance of an autobiographical memory task, while tolcapone was associated with increased function in frontal, parietal and striatal regions during the performance of an emotional working memory task.

**Conclusions:** This systematic review identified preliminary evidence for normalizing brain function after use of alternative pharmacological agents to first-line pharmacological treatments.

## Introduction

Posttraumatic stress disorder (PTSD) is a severe psychiatric disorder that can arise after exposure to a traumatic event. Although around 6% of all people exposed to a traumatic event will develop PTSD ^1–3^, these rates are significantly higher (25–35%) for assault victims, refugees, or combat veterans ^4^. Symptoms of PTSD include persistent re-experiencing of the traumatic event, avoidance of trauma-associated stimuli, negative thoughts or feelings, and increased arousal that persist for at least a month after trauma exposure ^5^. Clinical manifestations and symptoms intensity of PTSD are very heterogeneous ^6^, possibly implying the involvement of multiple neurobiological systems ^7^.

Among the numerous theories proposed to explain PTSD, including the emotional processing to fear model ^8^, the dual representation theory ^9^, or the cognitive model of PTSD ^10^, most neuroimaging studies have been based on the failure to inhibit fear model of PTSD ^11, 12^. This model implies insufficient top-down regulation from the medial prefrontal cortex to the amygdala ^11–13^, leading to hyperactivation of the amygdala. Changes in amygdala-medial prefrontal cortex connectivity can lead to failure to suppress the so-called default mode network ^14^ during cognitive engagement^15^. This aberrant top-down regulation can be normalized following successful psychological- and/or pharmacological treatments ^16^. Anatomical results from the Enhancing NeuroImaging Genetics through Meta-Analyses (ENIGMA) consortium indicate that smaller hippocampal and amygdala, in addition to smaller left and right lateral orbitofrontal gyri, were evident in people with current PTSD compared with trauma-exposed control subjects ^17, 18^; although smaller hippocampal volume may be present prior to trauma exposure and could represent a risk factor to develop PTSD following trauma exposure ^19, 20^. Despite evidence for increased medial prefrontal cortex activation following successful treatment, the effects of treatment on amygdala activation or volume remains unclear ^16, 21^. Other regions, such as the thalamus, the precuneus and the occipital cortex have also been inconsistently associated with PTSD and/or trauma exposure ^22–24^.

Both psychological and pharmacological approaches have been found effective for treating PTSD. First-line psychological interventions for PTSD include trauma-focused cognitive behavioural therapy, eye movement desensitization and reprocessing, or prolonged exposure therapies ^25^, while antidepressants such as selective serotonin reuptake inhibitors (SSRIs), tricyclic antidepressants, or monoamine oxidase inhibitors have been recommended as first-line pharmacological options ^7^. A meta-analysis of pharmacological treatments indicates that sertraline, fluoxetine, paroxetine, prazosin, venlafaxine, quetiapine, and risperidone, have a small positive effect with no evidence of superiority for one intervention over another ^26^. The use of SSRIs has been linked with both changes in brain function and morphology in PTSD ^16^: fluoxetine was found to normalize hyperactivity of the cerebellum, precuneus, and supplementary motor cortex ^27^, while paroxetine was associated with increased hippocampal volume ^28^ and increased frontal (orbitofrontal cortex, anterior cingulate cortex) function during trauma versus neutral script presentations ^29^.

Evidence for the efficacy of current medications is limited ^30^, with about one-third of PTSD patients still meeting criteria for PTSD after treatment, and alternative approaches have been proposed ^31^. Other pharmacological agents, such as oxytocin ^32^, were found to enhance neural responses during reward and punishment anticipation in reward pathway regions including the ventral tegmental area, striatum, and insula, in non-trauma exposed healthy individuals ^33, 34^, as well as to normalize functional connectivity between sub-regions of the amygdala with the prefrontal cortex to the level of healthy trauma-exposed controls ^35^. A meta-analysis proposed that early administration of hydrocortisone after a traumatic event could prevent the development of PTSD ^30^. Glucocorticoid administration may offer critical therapeutic advances ^36^, by reducing the retrieval of traumatic memories and promote extinction and inhibitory fear learning ^30^, and can enhance the effects of prolonged exposure therapy ^37^. Another recently used pharmacological agent used for treating PTSD includes the beta-blocker propranolol: promising results indicate that the use of propranolol can reduce the severity of PTSD symptoms, and improve cognitive performance in patients with PTSD^38, 39^.

A recent review evaluated the effects of successful psychotherapy on the brain in PTSD patients and reported that psychotherapy may up-regulate the medial prefrontal cortex, leading to symptom reduction ^40^. However, the neural correlates underlying successful pharmacological intervention in PTSD remain unclear. Understanding the effects of these new treatment options on brain function is critical to extend existing models, in order to consider the translational value of selected drugs or drug receptors into clinical therapy ^41^ and to provide the best options to people suffering from PTSD. The current study reviews the clinical trials investigating the functional brain changes associated with the use of these new pharmacological agents in PTSD patients. We aim to clarify the effects of these pharmacological agents on brain function in patients with PTSD. We hypothesise that these new pharmacological agents will normalise the dysregulated fronto-limbic function and connectivity.

## Materials and methods

### Search strategy

Adhering to the Preferred Reporting Items for Systematic Reviews and Meta-Analyses (PRISMA) 2020 statement ^42^, the PubMed, Scopus, and Web of Science databases were searched for studies published before September 2022. The search was limited to clinical trials estimating the effects of pharmacological agents other than first-line pharmacological treatments (e.g., SSRIs), on resting-state or task-related fMRI outcomes. The search was performed on PubMed using the following MESH terms: Stress Disorders, Post-Traumatic AND Magnetic Resonance Imaging. The search was then restricted to Clinical Trials by applying search filters “article type: Clinical Trial.” Search on Web of Science and Scopus using the search terms *MRI, fMRI, neuroimaging, magnetic resonance imaging* crossed with *therapeutic, treatment, intervention* and again fully crossed with the terms *posttraumatic stress disorder, PTSD*. The studies selection process is outlined in the flow diagram (see Figure 1).

**Figure 1.**
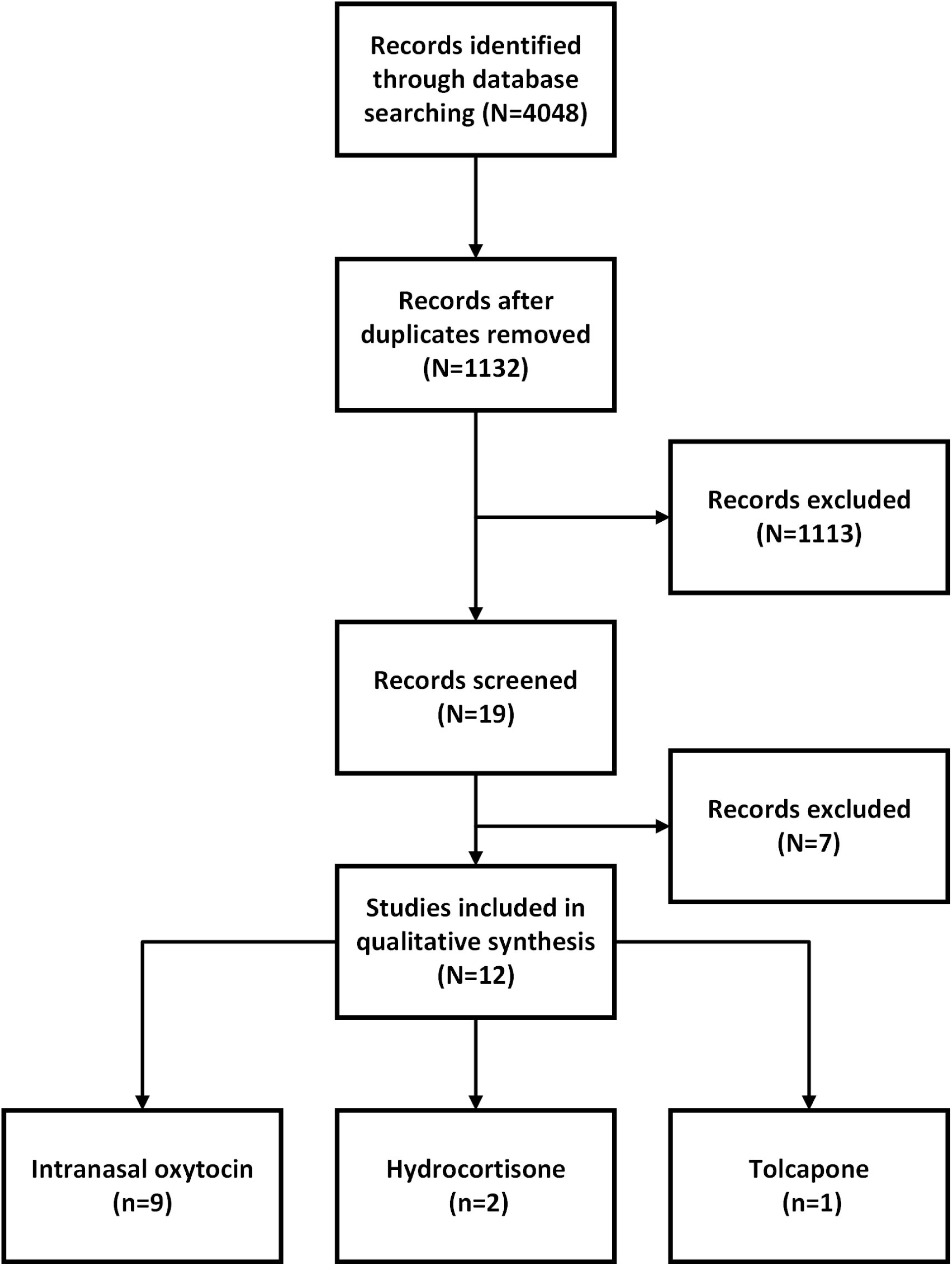
PRISMA Flow diagram

### Study selection

Inclusion criteria included original articles on adult patients with PTSD, diagnosed based on either the criteria from the Diagnostic and Statistical Manual of Mental Disorders fourth or fifth editions (DSM-IV, DSM-IV-TR, DSM-5) ^5, 43, 44^ or the International Statistical Classification of Diseases and Related Health Problems (ICD-10) ^45^. Studies including a healthy control group and/or a group of trauma-exposed controls (TEC) who did not develop PTSD following trauma exposure (healthy controls, HC) were considered. Studies that were not clinical trials, in which patients had severe comorbid disorders, studies that did not report fMRI as their outcome measures, studies with incomplete reporting, studies comparing two pharmacological interventions, studies not including an HC or TEC comparison groups, and studies published in other languages than English were excluded. Two reviewers independently screened the identified abstracts. Extracted data included the year of publication, sex, age, diagnosing tool, type of fMRI (resting state of task-related fMRI), and the name of the task for task-related fMRI studies.

### Data extraction

Study characteristics were extracted, including sample size, diagnostic tools, type of treatment and type of brain imaging (see Table 1). In addition, the relevant results from each study, including the main effects of the treatment of interest, the main effects of groups and the effects of the group-by-treatment interactions on brain function were extracted and reported in Table 2. Other results within each study were not reported here if they were not related to the aims of the present study.

**Table 1.**
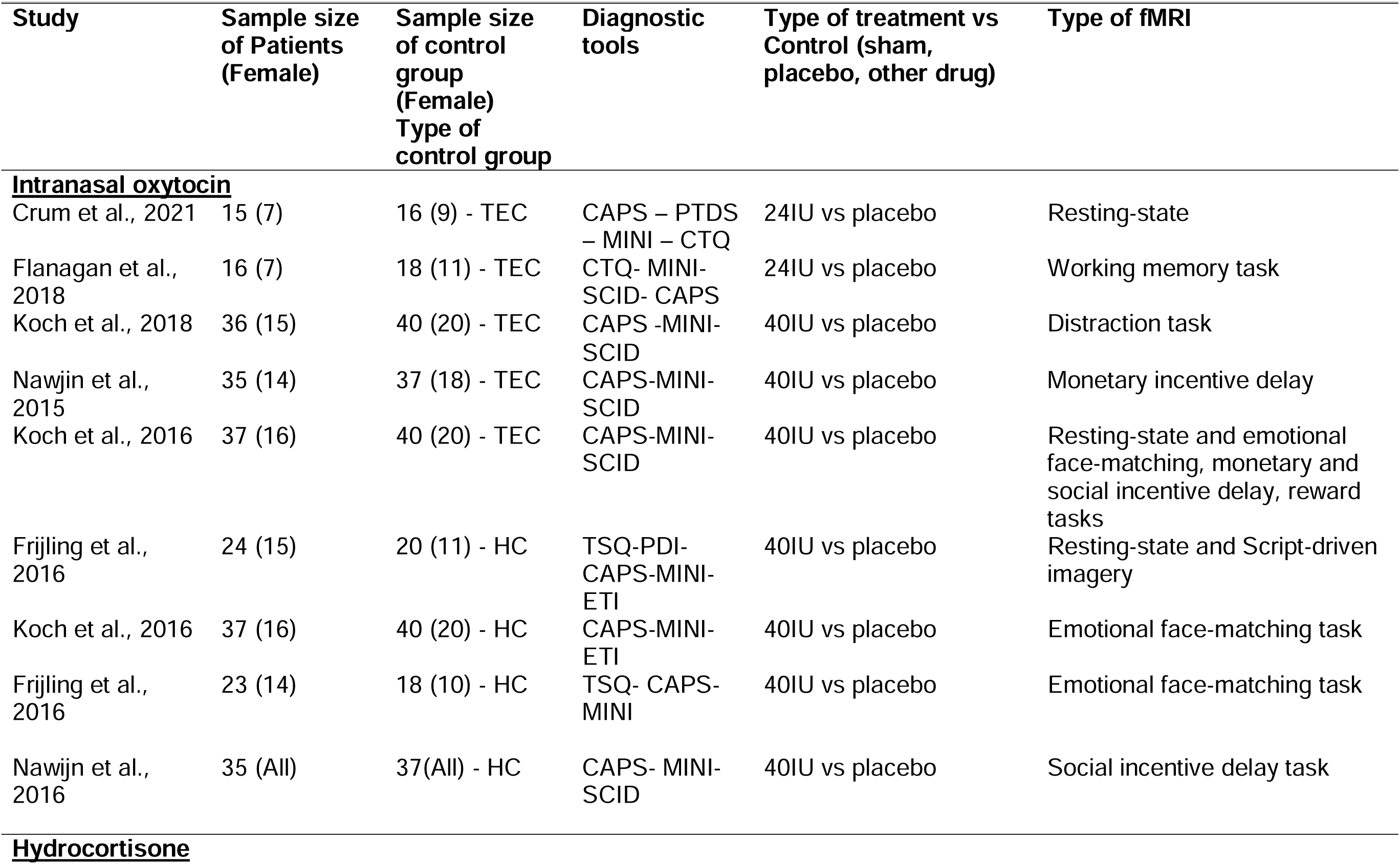
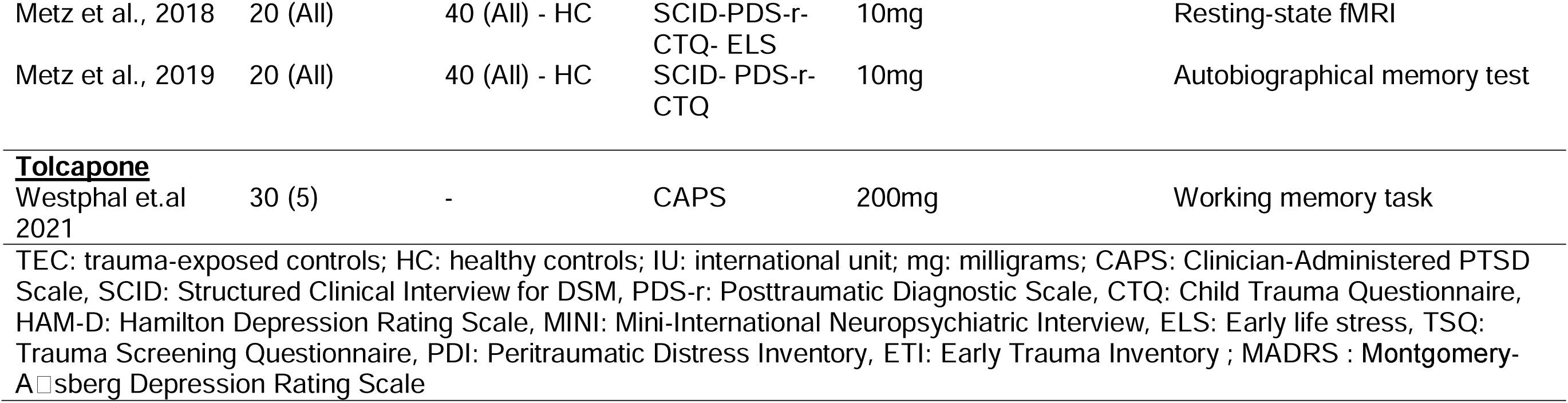
Details of the included studies

| Study | Sample size of Patients (Female) | Sample size of control group (Female)<br>Type of control group | Diagnostic tools | Type of treatment vs Control (sham, placebo, other drug) | Type of fMRI |
| --- | --- | --- | --- | --- | --- |
| **<u>Intranasal oxytocin</u>** |  |  |  |  |  |
| Crum et al., 2021 | 15 (7) | 16 (9) - TEC | CAPS – PTDS – MINI – CTQ | 24IU vs placebo | Resting-state |
| Flanagan et al., 2018 | 16 (7) | 18 (11) - TEC | CTQ- MINI- SCID- CAPS | 24IU vs placebo | Working memory task |
| Koch et al., 2018 | 36 (15) | 40 (20) - TEC | CAPS -MINI- SCID | 40IU vs placebo | Distraction task |
| Nawjin et al., 2015 | 35 (14) | 37 (18) - TEC | CAPS-MINI- SCID | 40IU vs placebo | Monetary incentive delay |
| Koch et al., 2016 | 37 (16) | 40 (20) - TEC | CAPS-MINI- SCID | 40IU vs placebo | Resting-state and emotional face-matching, monetary and social incentive delay, reward tasks |
| Frijling et al., 2016 | 24 (15) | 20 (11) - HC | TSQ-PDI- CAPS-MINI- ETI | 40IU vs placebo | Resting-state and Script-driven imagery |
| Koch et al., 2016 | 37 (16) | 40 (20) - HC | CAPS-MINI- ETI | 40IU vs placebo | Emotional face-matching task |
| Frijling et al., 2016 | 23 (14) | 18 (10) - HC | TSQ- CAPS- MINI | 40IU vs placebo | Emotional face-matching task |
| Nawijn et al., 2016 | 35 (All) | 37(All) - HC | CAPS- MINI- SCID | 40IU vs placebo | Social incentive delay task |
| **<u>Hydrocortisone</u>** |  |  |  |  |  |
| Metz et al., 2018 | 20 (All) | 40 (All) - HC | SCID-PDS-r-CTQ- ELS | 10mg | Resting-state fMRI |
| Metz et al., 2019 | 20 (All) | 40 (All) - HC | SCID- PDS-r-<br>CTQ | 10mg | Autobiographical memory test |
| **Tolcapone** |  |  |  |  |  |
| Westphal et.al<br>2021 | 30 (5) | - | CAPS | 200mg | Working memory task |

TEC: trauma-exposed controls; HC: healthy controls; IU: international unit; mg: milligrams; CAPS: Clinician-Administered PTSD Scale, SCID: Structured Clinical Interview for DSM, PDS-r: Posttraumatic Diagnostic Scale, CTQ: Child Trauma Questionnaire, HAM-D: Hamilton Depression Rating Scale, MINI: Mini-International Neuropsychiatric Interview, ELS: Early life stress, TSQ: Trauma Screening Questionnaire, PDI: Peritraumatic Distress Inventory, ETI: Early Trauma Inventory ; MADRS : Montgomery-Åsberg Depression Rating Scale

**Table 2.**
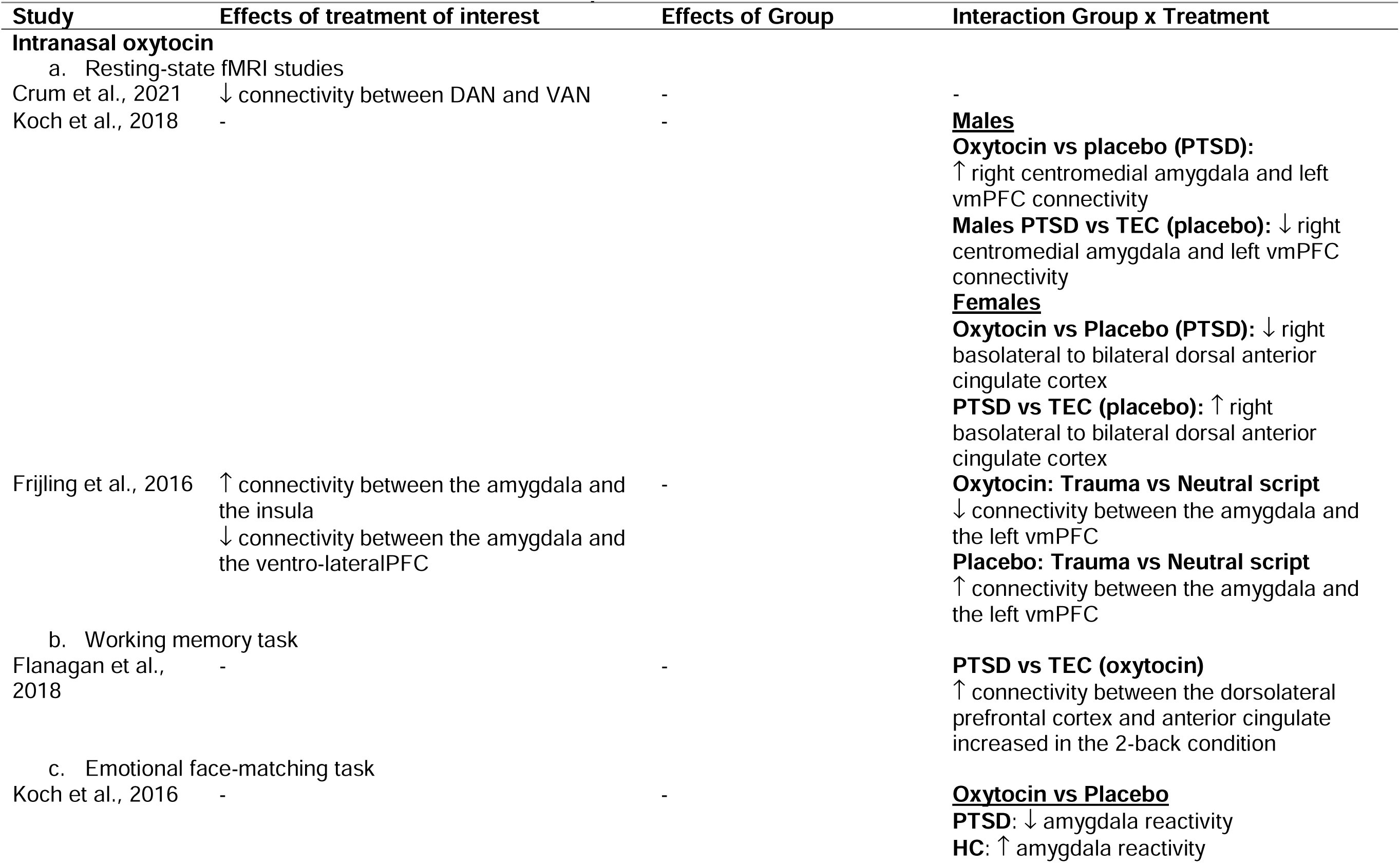
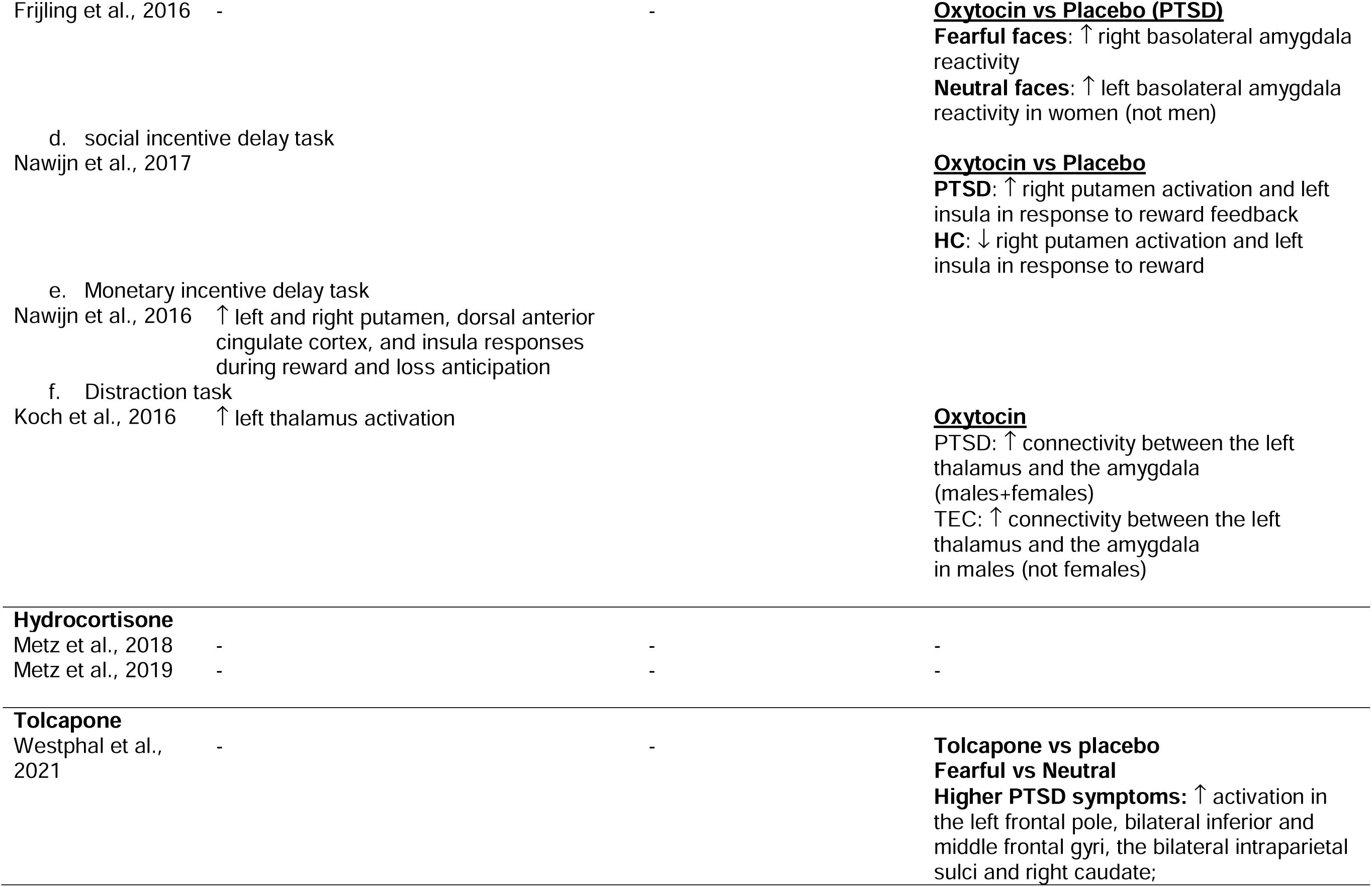
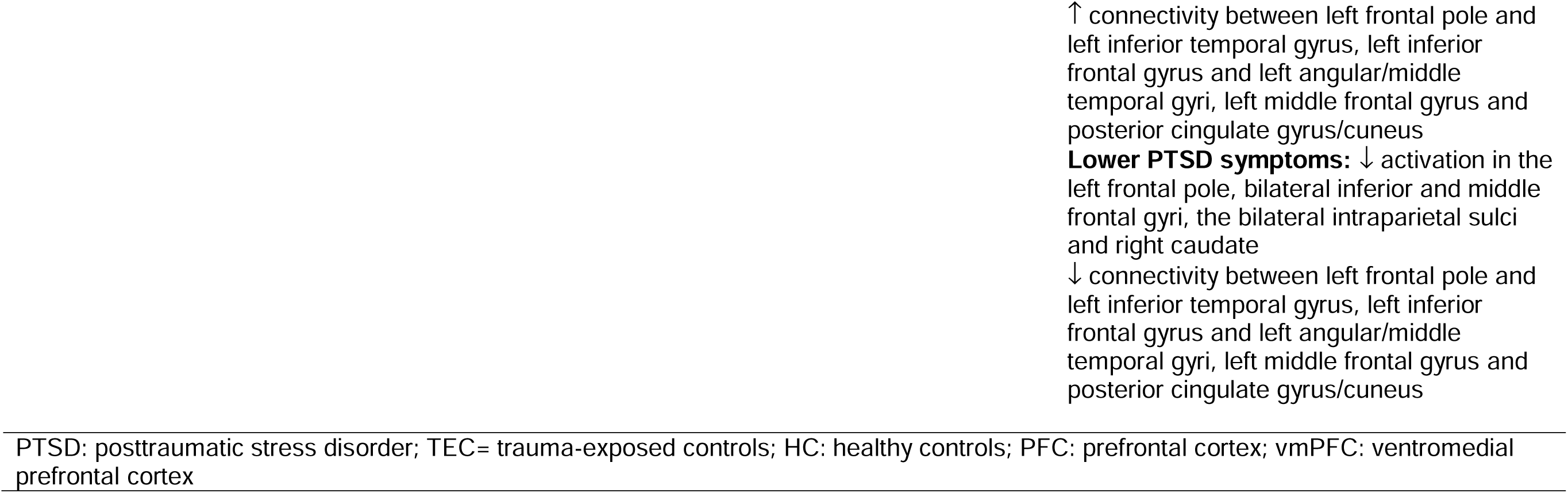
Effects of interest extracted from the included publications

### Risk of bias assessment

Two authors (SL and AY) independently assessed the risks of bias using the Cochrane Collaboration’s Tool for Assessing Risk of Bias in Randomized Trials (RoB 2.0) ^46^. In addition to provide an overall bias estimation, this tool assesses five domains of potential bias for each individual study (selection of the reported result, measurement of the outcome, missing outcome data, deviation from intended interventions and randomization process), and ranks them as ‘low’, with ‘some concern’ or ‘high’. In the event of discrepancy in bias assessment between reviewers, consensus was achieved through discussion.

## Results

A total of 4,048 articles were identified. After removing duplicates, abstracts and titles for 1,132 papers were screened, from which 1,113 articles were excluded. Finally, seven articles were removed because of highly comorbid disorders with PTSD, and 12 articles were included in the review (see Figure 1 and Table1). Among the 12 included studies investigating the changes in brain function after administration of pharmacological treatment in patients with PTSD, nine used intranasal oxytocin (3 resting-state fMRI, 6 task-related fMRI), two used hydrocortisone (1 resting-state fMRI, 2 task-related fMRI), and one task-related fMRI used Tolcapone. Effects of treatments, groups and their interactions on brain function are reported in Table 2.

### Oxytocin

#### Resting-state fMRI

Koch et al. (2016) used resting-state fMRI to investigate the effects of oxytocin on functional connectivity of the basolateral and centromedial amygdala in police officers with (n=37) and without (n=40) PTSD ^35^. Follow-up post-hoc analyses were conducted separately in males and females, showing that the connectivity between the right centromedial amygdala and the left ventromedial prefrontal cortex (vmPFC) was normalised (enhanced) in response to oxytocin administration in male PTSD cases relative to placebo, reaching a similar level than the TEC group after oxytocin administration. Connectivity between these two regions was reduced in PTSD males compared to TEC males after placebo. There were effects of intervention or group on connectivity between left ventromedial prefrontal cortex and right centromedial amygdala in females. However, placebo was associated with enhanced connectivity between the right basolateral to bilateral dorsal anterior cingulate cortex in PTSD females, compared to TEC females. In addition, PTSD females showed reduced connectivity between these regions following oxytocin administration, compared to placebo ^35^.

In another study of civilian acute PTSD cases recruited early following trauma exposure (scanning session within 11 days post trauma) ^47^, functional connectivity at rest between the (whole) amygdala and the left ventrolateral prefrontal cortex was weaker in response to a previously presented trauma-related script compared to a neutral script following oxytocin administration, while this association was stronger in the placebo group. Irrespective of the script condition, administration of oxytocin increased the strength of functional connectivity between the amygdala and the insula, and decreased the strength of connectivity between the amygdala and the vmPFC ^47^.

Finally, oxytocin was found to reduce the strength of resting state connectivity between the dorsal (DAN; made of the precuneus, occipital cortex, parietal cortices, middle and superior frontal gyri, temporooccipital gyrus, and precentral gyrus) and ventral attentional networks (VAN; made of the supplemental motor area/superior frontal gyrus, bilateral superior and middle temporal gyrus, and bilateral inferior frontal gyrus) in both a group of cases with childhood trauma related PTSD and TEC ^48^. In this study there was no specific effects of oxytocin in the PTSD group.

#### Working memory

Only one study investigated the effects of oxytocin on brain function during the performance of an n-back working memory task in people with PTSD relative to childhood trauma exposure compared to childhood trauma TEC ^49^. Compared to placebo, in the context of improved performance at the most difficult condition (2-back), connectivity between the dorsolateral prefrontal cortex and anterior cingulate cortex increased in the 2-back condition among individuals with PTSD using oxytocin (relative to TEC) ^49^.

#### Emotional face-matching

Two studies have investigated the effects of intranasal oxytocin administration while performing an emotional face-matching task. In their cohort of police officers, Koch et al. (2016) also examined the effects of oxytocin on amygdala reactivity toward emotional stimuli ^50^. Compared to placebo, oxytocin administration was associated with decreased amygdala reactivity in PTSD patients and enhanced amygdala reactivity in healthy controls. The intervention moderated the relationship between pre-intervention anxiety scores and amygdala reactivity: this association between anxiety and amygdala reactivity was dampened after oxytocin administration, while increased in the placebo group. In their cohort of acute civilian PTSD patients, a single intranasal oxytocin administration showed enhanced right basolateral amygdala reactivity to fearful faces compared to the placebo group. In addition, enhanced reactivity of the basolateral amygdala in response to neutral emotions (but not happy or fearful) was found in women (but not men) using oxytocin compared to placebo ^51^.

#### Social incentive delay task

Only one study investigated brain response to oxytocin when performing a social incentive delay task in police officers ^52^. Compared to placebo, intranasal oxytocin administration was associated with increased activation in the right putamen and left insula in response to reward feedback in the PTSD group, while decreased activation in these regions was evident in the control group.

#### Monetary incentive delay task

Only one study evaluated the effects of oxytocin on brain function during the performance of a monetary incentive delay task in police officers with or without PTSD ^53^. Compared to placebo, oxytocin administration enhanced neural responses during reward and loss anticipation in both PTSD patients and controls in the left and right putamen, dorsal anterior cingulate cortex, and insula. Reward processing in the ventral striatum was also positively associated with anhedonia after oxytocin administration, suggesting that oxytocin may increase motivation.

#### Distraction task

The only study investigating the impact of oxytocin administration on brain response during the performance of a distraction task ^54^, reported that former police officers showed enhanced left thalamus activation during the task after oxytocin administration compared to placebo, independently of being diagnosed or not with PTSD. In addition, oxytocin administration was associated with increased task-related functional connectivity between the left thalamus and the amygdala in all PTSD patients and in male, but not female, TEC.

Overall, administration of oxytocin was associated with normalisation of functional connectivity, at rest or during the performance of a cognitive task, and patterns of activation in critical regions involved in PTSD (e.g., amygdala-ventral prefrontal cortex at rest, dorsolateral prefrontal and anterior cingulate cortices during working memory).

### Hydrocortisone

One group studied the effects of hydrocortisone on brain function of females with PTSD, borderline personality disorder and healthy controls. Results reported no effects of hydrocortisone, or placebo, on patterns of resting-state functional connectivity ^55^ or brain function during the performance of an autobiographical memory task ^56^.

### Tolcapone

Only one study investigated the effects of tolcapone, a nitrocatechol-type inhibitor of the enzyme catechol-O-methyltransferase, on brain function during the performance of an emotional working memory task in 30 veterans ^57^. Results revealed a significant association between a three-way interaction between intervention (tolcapone vs placebo), PTSD severity and the stimulus emotion (neutral vs fearful) and brain function. In particular, results indicated that increased activation in bilateral middle frontal gyri, the left frontal pole, bilateral inferior and the bilateral intraparietal sulci and right caudate (from region-of-interest analysis) in response to fearful stimuli (versus neutral) was evident following tolcapone administration (compared to placebo) in PTSD patients reporting higher levels of PTSD severity (both at one and two standard deviations above the average PTSD severity). On the other hand, decreased activation was evident for those reporting lower levels of PTSD symptoms (one standard deviation below the average PTSD severity). Similar patterns of task-related functional connectivity were reported between frontal seed regions (left frontal pole, left inferior frontal gyrus, left middle frontal gyrus) and the left inferior temporal gyrus, left angular/middle temporal gyri, and posterior cingulate gyrus/cuneus, respectively.

### Bias assessment

The overall quality assessment of the selected articles was low, with 60% of studies included showing some concerns and 40% being considered at high risk (see Figure 2). Notably, the deviations from intended interventions were estimated being at low risk in only 10% of the selected studies (50% with some concerns, 40% at high risk), while the selection of reported results was estimated to be at low risk for 50% of the included studies (40% with some concerns and 10% at high risk), and the measurement of the outcome and the missing outcome data were at low risk in 40% (20% with some concerns, 40% at high risk for the measurement of the outcome; 30% with some concerns and 30% at high risk for the missing outcome). Finally, the randomization process was estimated being at low risk of bias in 90% of the included studies (10% with some concerns).

**Figure 2.**
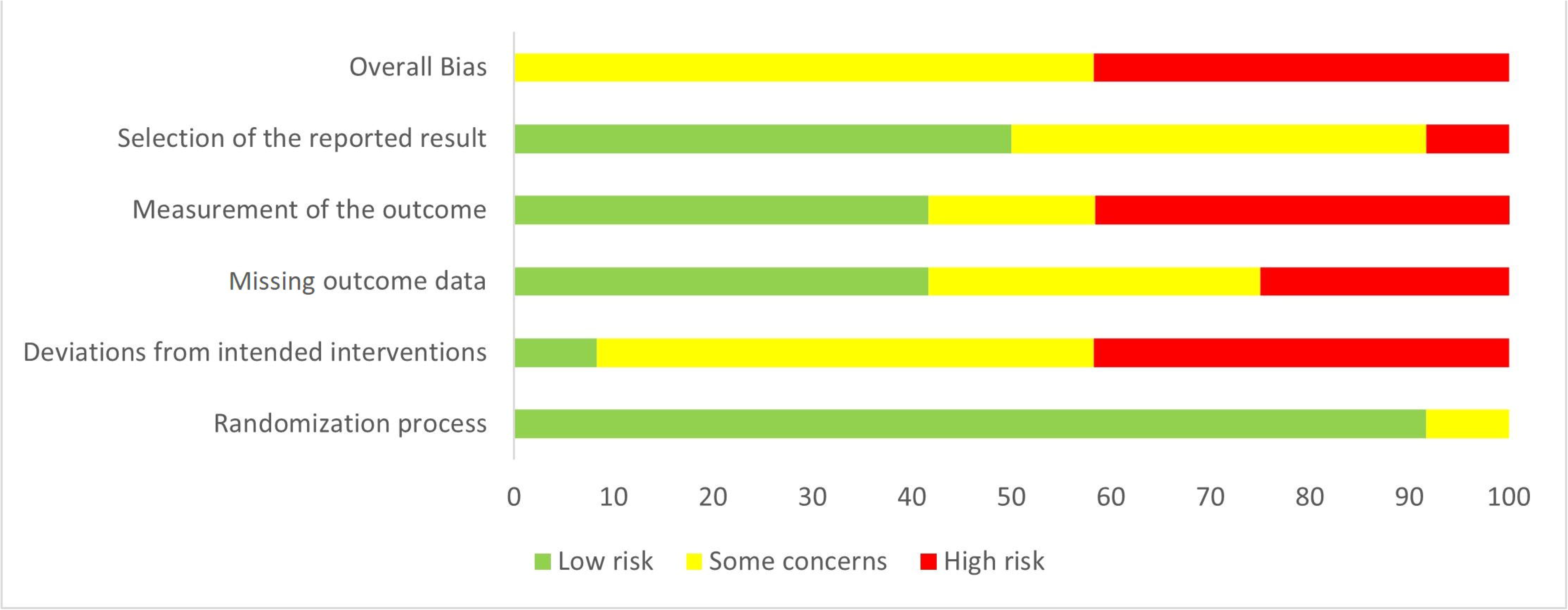
Bias assessment of the included studies

## Discussion

The present systematic review identified 12 clinical trials investigating the impacts of pharmacological agents (oxytocin, hydrocortisone, tolcapone) on brain function in PTSD. Although these studies used various imaging paradigms (resting-state fMRI, task-related fMRI paradigms), results mostly converge toward functional changes in the prefrontal cortex, cingulate cortex, amygdala, cerebellum, parietal cortex.

### Functional brain changes after oxytocin administration

At rest, oxytocin administration was associated with changes in functional connectivity between frontal regions (e.g., vmPFC) and limbic and affective regions (e.g., amygdala). Normalisation (increase) of the connectivity between the vmPFC and the amygdala, especially its centromedial nucleus, was evident in male, but not female, patients with PTSD ^35^. This was further evident following exposure to a trauma-related script ^47^. Based on the amygdalo-centric model of PTSD ^12^, this increased connectivity may reflect increased top-down prefrontal control over the fear response. Interestingly, oxytocin administration was also associated with reduced connectivity strength between dorsal (mostly cognitive) and ventral (mostly affective) attentional networks ^48^, suggesting that this pharmacological agent may help reduce focus on affective inner thoughts.

During the performance of cognitive tasks, oxytocin administration was associated with changes in activation in brain regions critical for the tasks studied. During the performance of a working memory task, oxytocin administration was associated with, increased connectivity between, the left dorsolateral prefrontal and anterior cingulate cortices, with greater effects in people diagnosed with PTSD compared to TEC ^49^. The direction of effect observed between these two regions was not consistent with normalisation of the aberrant patterns of connectivity in PTSD, suggesting that oxytocin-related enhanced recruitment of this circuit may reflect some compensatory mechanisms to adequately perform the task, and may not necessarily reflect increased cognitive efficiency or performance monitoring. During the performance of an emotional face matching task, results were inconsistent: administration of oxytocin (compared to placebo) was associated with decreased amygdala activity in a group of police officers ^50^, while associated with increased reactivity of the right basolateral amygdala in a group of acute civilian PTSD patients ^51^. This discrepancy may be related to the fact that police officers may have been repeatedly exposed to stressful, if not traumatic events, than the group of acute PTSD patients. Sensitization of the stress system to repeated stress exposure may impact the baseline levels of amygdala reactivity and influence the way oxytocin modulates it. This explanation remains speculative and future studies including groups of PTSD patients with different levels of basal stress and trauma exposure may help understanding how oxytocin administration may differently influence emotional processing. This is critical to provide the most efficient intervention to people suffering from PTSD.

Other studies have reported enhanced activation in striatal regions following oxytocin administration when performing rewarding tasks, such as social or monetary incentive delay task ^52, 53^. While oxytocin was associated with increased putamen, anterior cingulate and insular activation independently of the group during reward and loss anticipation, increased putamen and insular activations were specifically observed in PTSD, and decreased in HCs, when social reward feedback was sought after. These findings may indicate that reward evaluation in PTSD may be facilitated by oxytocin administration that may be able to enhance motivational processing. However, the impact of oxytocin on brain function during these tasks is highly sensitive to sex, baseline levels of oxytocin system function, and potentially type of traumatic event and chronicity of PTSD ^58^. Finally, when performing a distraction task ^54^, oxytocin administration was found to modulate thalamic activation, and connectivity with the amygdala, suggesting that oxytocin administration can enhance neural emotional control in PTSD. As suggested by the authors, intranasal oxytocin administration could be used as an enhancer for the changes in brain function produced by first-line psychological approach for PTSD, such as cognitive-behavioural therapy. This will need to be further tested in future trials. If these effects are confirmed, the association of oxytocin and CBT may provide important new avenues for the treatment of PTSD.

### Functional changes after hydrocortisone and tolcapone administration

Effects of other pharmacological agents in PTSD are less evident. For instance, hydrocortisone administration was not associated with any changes in brain function, either at rest or during the performance of an autobiographical memory task ^55, 56^. The relatively small sample size of these studies may have limited the power to detect more subtle effects. It may also be the case that hydrocortisone may have long-term effects, not captured in the present trials. It would be important to replicate and confirm the lack of effects in larger populations, potentially in groups of patients exposed to different types of traumas. On the other hand, administration of tolcapone, a nitrocatechol-type inhibitor of the enzyme catechol-O-methyltransferase, was associated with increased activation and connectivity during the performance of an emotional working memory task, among regions critical for working memory (e.g., middle frontal gyrus, parietal lobules and caudate) and social cognitive processes (e.g., frontal pole, inferior frontal gyrus, temporo-parietal junctions, posterior cingulate gyrus) in PTSD patients reporting larger PTSD symptoms, that was reduced in those reporting less severe PTSD symptoms ^57^. These effects suggest that tolcapone may help reducing the emotional distraction experienced by people with severe PTSD symptoms. On the other hand, increasing dopamine following tolcapone in those with less severe PTSD symptoms, may have influenced emotional working memory processing, similar to the effects generally observed in stressful situations. However, these results need replication and extension.

### Limitations

This systematic review has several limitations. First, we only focused on fMRI studies, excluding alternative methods to investigate neural alterations (e.g., positron emission tomography, single-photon emission computerized tomography, electro- encephalography). In addition, the interpretation of the findings mentioned above are limited by the heterogeneous research methods used, including the types of tasks, as well as the way drugs were administered (e.g., intranasal versus oral administration). Other recent studies using ketamine ^59, 60^ or psychedelics ^61^ were not included as they did not compared patients with PTSD to groups of TEC or HCs. An important limitation is the exclusion of studies of PTSD patients showing severe comorbid disorders; although this would represent a more ecological group, it could also hinder the effects of the studied pharmacological agents on brain function solely associated with PTSD. Furthermore, symptoms severity, differences in trauma types and age of trauma onset were not considered. Finally, the included studies were in general of small sample sizes, limiting their replicability and generalizability.

## Conclusions

Despite the heterogeneity of methods and pharmacological agent used, the present systematic review of clinical trials identified preliminary evidence for normalization of brain function following the administration of pharmacological agents, including intranasal oxytocin, hydrocortisone and tolcapone. Overall, normalisation of the vmPFC-amygdala connectivity following oxytocin administration was the most consistent finding, but interpretation of the results from this systematic review are limited by the limited number of clinical trials included. Further clinical trials are necessary to better understand the neurobiological mechanisms and effects of these pharmacological agents on brain function and morphology, and their benefits in helping people suffering from PTSD.

## Data Availability

The data that support the findings of this study are available from the corresponding author, upon reasonable request.

## Ethics approval and consent to participate

No informed consent or ethical approval is required for the purpose of this review.

